# Dietary sugar exposure in early life and risk of adult mental health disorders: UK Biobank cohort study

**DOI:** 10.64898/2026.01.20.26344391

**Authors:** Hana F Navratilova, Anthony D Whetton, Nophar Geifman

## Abstract

Recent findings suggest that sugar rationing during the first 1000 days of life may influence chronic disease risk in adulthood. Given the link between mental health and non-communicable diseases, early-life sugar restriction may be associated with later-life incidence of anxiety and depression. Using UK Biobank data, participants born between October 1, 1951, and March 31, 1956 (n = 46,448) were grouped by duration of exposure to sugar rationing: R270 (rationed from conception to 270 days of life, n = 9,671), R425 (n = 6,984), R635 (n = 6,622), R817 (n = 7,096), R1000 (n = 6,856), and never-rationed groups NR0 (conceived immediately after sugar rationing ended, n = 6,117) and NR455 (conceived after all food rationing ended, n = 3,102). The liking of sweet foods was assessed via food preference questionnaires, and anxiety/depression diagnoses via linked ICD-10 health records. Survival analyses were conducted using Weibull accelerated failure time models, adjusted for demographic, lifestyle, and health covariates. Structural equation modelling (SEM) was used to assess outcomes and causal pathways. The R1000 group reported lower sweets preference (mean score = 5.82 vs. 6.04 in NR455, p = 0.0042) and showed significantly reduced hazards of anxiety (HR = 0.67, 95% CI: 0.53–0.84) and reduced hazards of depression (HR = 0.71, 95% CI: 0.57–0.87). Protective associations for anxiety remained robust across sensitivity analyses, including adjustment for later-life sugar intake, whereas associations for depression were attenuated and non-significant. SEM confirmed a direct effect of sugar rationing duration on anxiety (γ = – 0.144, p = 0.012), but not depression. Exposure to sugar rationing during the first 1000 days of life was associated with reduced risk of anxiety in adulthood, independent of later dietary behaviour. These findings highlight the importance of early developmental environments in shaping psychiatric outcomes and suggest that nutritional exposures may confer resilience against anxiety disorders. Effects on depression were weaker and appear partly explained by later-life factors.

## INTRODUCTION

Numerous studies have thoroughly examined the link between quality of diet and human health, revealing that diet influences not only metabolic diseases but also the risk of mental health conditions [1–3]. Among various dietary components, sugar consumption has been particularly scrutinised for its potential impact on mental well-being across different populations. High sugar intake has been linked to an increased risk of depression and anxiety [4–8], underscoring the importance of understanding how dietary choices influence mental health. Moreover, a higher liking for sweet foods has also been found to be associated with depression and anxiety [9]. This is important because food liking can often serve as a proxy for actual intake; individuals who have a higher preference for sweet foods are more likely to consume them in greater quantities [10], potentially exacerbating the risk of developing related health issues or lowering the adherence to interventions such as controlled diets.

Depression and anxiety are multifactorial diseases linked to biological, genetic, and environmental factors, with epigenetic mechanisms potentially mediating increased depression risk after adverse life events [11]. Recent findings from the United Kingdom (UK) suggest that sugar rationing during the first 1000 days of life may contribute to the risk of chronic diseases, particularly diabetes, cardiovascular diseases, and hypertension [12–15]. Given that depression and anxiety are often associated with other mental conditions and non-communicable diseases [16–18], early-life sugar rationing might also contribute to the incidence of these mental health issues later in life.

Sugar rationing in the UK, implemented to manage severe food shortages caused by World War II, lasted from January 1940 to September 1953, limiting adults to approximately 40 grams of sugar per day [19]. The UK Biobank, which holds extensive health and lifestyle data on over 500,000 older adults in the UK, provides a valuable resource for examining the long-term effects of early-life conditions. Over half of UK Biobank participants were born during the sugar rationing period, with birth years spanning from the early 1940s to the early 1970s, enabling unique analyses of early-life sugar exposure by comparing those who had no exposure to sugar during critical developmental stages under rationing with those born after it ended and when sugar was reintroduced into diets.

This study aims to explore the interconnections between early-life sugar exposure, liking for sweet foods, sugar intake, and mental health risks, particularly depression and anxiety. We investigate whether early-life sugar rationing is associated with sugar preferences in later life and examine the potential links between sugar exposure in early life and mental health outcomes. Depression and anxiety are now understood to be multifactorial disorders with complex genetic, environmental, and developmental influences [20]. Inflammation has been linked to both depression and anxiety [21–24], as has structural brain alterations and altered connectivity evident in major depressive disorder [24–26]. These potential mediators of risk, as opposed to others, can be, to some extent, affected by nutritional exposures. Thus, this study also considers inflammatory levels and grey matter volume as possible alternative pathways. By addressing these gaps, this research could provide valuable insights into preventive strategies and dietary recommendations for improving mental health.

## METHOD

### Study Population

This study used data from the UK Biobank (accessed under application number 83988). The UK Biobank recruited more than 500,000 people aged 37–73 years from the United Kingdom between 2006 and 2010. The data consisted of a wide range of phenotypic information and biological samples, including demographic characteristics, mental health, blood assays, multimodal neuroimaging, and other parameters. Participants born between October 1, 1951, and June 30, 1954 were identified to represent those who were conceived and born within the period of sugar rationing, as previously described [12]. Comparator never-rationed groups who had exposure to sugar from conception consisted of individuals born immediately after the cessation of sugar rationing (July–December 1954) and those born a year after the end of all food rationing (January–March 1956). Exclusion criteria included birth or residence outside the UK, multiple births, adoption, withdrawal from the Biobank, and pregnancy at the time of survey. The final analytic sample comprised 46,448 participants.

Rationed group participants were categorised as follows: R270, born between October 1953 and June 1954, exposed to sugar rationing in-utero only, for a duration of approximately 270 days (n=9671); R425, born between April 1953 and September 1953, rationed in-utero and up to 6 months of age, for approximately 425 days (n=6984); R635, born between October 1952 and March 1953, rationed in-utero and up to 12 months of age, for approximately 635 days (n=6622); R817, born between April 1952 and September 1952, exposed to sugar rationing in-utero and up to 18 months of age, for approximately 817 days (n=7096); and R1000, born between October 1951 and March 1952, rationed in-utero and up to 24 months of age, for approximately 1000 days (n=6856). Two comparison groups were defined as never-rationed: NR0 (July–December 1954, conceived immediately after sugar rationing ended, n=6117) and NR455 (January–March 1956, conceived after all food rationing ended, n=3102).

### Analysis of sweet food-liking, actual sugar consumption, and sugar rationing exposure

To examine the relationships between rationing status and preferences for sweet foods, free sugar intake, and total sugar intake, we calculated average scores for sweet food liking (Field ID 20732) and sugar consumption for a subset of participants (n = 15,058) who completed the Oxford WebQ, a self-administered 24-hour recall questionnaire, at least once, and also completed the Food Preference Questionnaire. The sugar intake data were averaged across 1-5 repeated 24-hour recall assessments. To gain a further understanding of the potential associations with liking of sweet foods, several specific sweet food preferences were examined; these include liking for: biscuits (Field ID 20615), cake (Field ID 20631), cake icing (Field ID 20632), croissants (Field ID 20648), dark chocolate (Field ID 20652), cheesecake (Field ID 20636), jam (Field ID 20680), marzipan (Field ID 2688), and milk chocolate (Field ID 20691). The selection of these specific items was based on our previous work [9].

### Health outcome assessment

Linked health outcome data (earliest record date and respective health outcome defined with ICD-10), obtained from UK Biobank, were sourced from primary care, inpatient hospital, and death record data. Self-reported data were not included in the analysis. Here, the primary outcome of anxiety was defined from ICD-10 codes F40 and F41 (Field ID 130904 and 130907), while depression status was defined from ICD-10 codes F32 and F33 (Field ID 130895 and 130896). Absolute risk was calculated by determining the number of individuals in each group who experienced anxiety or depression. This ratio, expressed as a percentage, represents the absolute risk. Two negative control diseases were also examined: type 1 diabetes mellitus (ICD-10 codes E10, E13, and E14) and post-traumatic stress disorder (ICD-10 codes F43). These conditions were selected based on their lack of direct biological or epidemiological links to sugar intake, age-related metabolic processes, or dietary exposures during early development.

### Structural Brain Imaging using MRI scan data

We obtained volumetric imaging data from the UK Biobank [27,28]. The number of participants with brain imaging data was 7,630. For the dependent variables, we used preprocessed T1-weighted structural MRI image-derived phenotypes, which included numerical volume data for 139 regional grey matter volumes (FAST) obtained from the UK Biobank imaging-processing pipelines. To correct for head size and total brain volume, we also used pre-processed T1-weighted image-derived phenotype numerical volume data for total white and grey matter, normalised for head size. Full scanning information is available in the UK Biobank Brain Imaging Documentation (biobank.ctsu.ox.ac.uk/crystal/docs/brain_mri.pdf).

### Statistical analyses

#### Survival Modelling

All analyses were conducted in R version 4.3.2. The incidence of anxiety and depression was modelled using accelerated failure time (AFT) survival regression with a Weibull distribution, providing hazard ratios (HRs) and 95% confidence intervals. Cases were right-censored at the last date of available ICD-10 hospital inpatient data from UK Biobank (30 September 2024), ensuring consistent follow-up across participants. Models were adjusted for time-invariant potential covariates: sex (female, male), Townsend deprivation index (quintiles), ethnic background (white and other), family history of mental health (yes/no), breastfeeding status (yes/no), birth month, and survey year. Additional covariates were included to account for variability in health status and lifestyle: BMI class (<18.5, 18.5–<25, 25–<30, 30–<35, 35–<40, ≥40 kg/m^2^), alcohol intake frequency (daily or almost daily, never, once or twice a week, one to three times a month, special occasions only, three or four times a week), household size (1, 2–4, ≥5), smoking status (never/previous/current), sleep duration (<7 hours, 7–<9 hours, ≥9 hours), and inflammation status (CRP <8.0 mg/dL, CRP ≥8.0 mg/dL).

In addition to survival modelling, descriptive analyses of the age at event were conducted. Among participants who experienced depression, the median age at diagnosis was calculated within each rationing group by aggregating the time-to-event variable restricted to cases with an event indicator. Kernel density estimates of age at event were also computed for each rationing group to characterise the distribution of onset ages. These descriptive measures provided complementary information to the regression models by summarising central tendency and distributional patterns of age at diagnosis across groups

#### Robustness checks

Robustness checks were conducted to evaluate the stability of findings. First, a placebo test was performed using the NRIR group, comprising of adults born in Ireland during the same period of time as those in the R1000 and R817 groups (born October 1951 – September 1952, n = 154). Although Ireland did experience sugar rationing, it ended earlier, in December 1951. Thus, Irish births overlapping with R1000 were exposed to rationing in-utero but not postnatally, while those overlapping with R817 were never rationed. This distinction allowed the NRIR group to serve as an external comparator, sharing birthdate overlap with UK cohorts but differing in rationing exposure. Second, missing data for categorical covariates were handled using multiple imputation by chained equations (MICE), implemented with the mice package[29] in R (five imputations, 10 iterations), with tailored imputation methods applied according to variable type and number of levels (e.g., logistic regression for binary variables, polytomous regression for unordered categorical variables, and proportional odds models for ordered factors). Third, interaction analyses were performed to assess gender modification of associations, with stratified models used where significant interactions were detected. Finally, later-life dietary exposures (average free sugar and total sugar intake) were incorporated in sensitivity analyses restricted to individuals with complete sugar intake data (n = 21,221) to evaluate whether adult diet influenced associations between rationing exposure and mental health outcomes.

#### One-way ANCOVA and post hoc tests

To compare measures of interest among the seven comparative (duration of rationing) groups, we conducted one-way ANCOVA analyses on brain MRI data traits. Levene’s test were employed to evaluate the homogeneity of variances prior to performing the ANCOVA. Subsequently, two-sample t-tests were conducted as post hoc analyses to investigate the differences among the seven groups. To ensure the accuracy of our results, standard covariates such as age, sex, BMI, and the Townsend deprivation index were included. Additional covariates, including brain volume normalised for head size, were also included to account for potential confounding factors. To correct for multiple comparisons, FDR corrections were applied.

#### Structural Equation Modelling (SEM)

A Structural Equation Model (SEM) was used to explore the relationships between duration of sugar rationing, inflammation levels, mental health outcomes (anxiety and depression), and three latent variables: preference for sweetness, sugar consumption, and brain MRI traits. For this analysis, we included only participants in the rationing groups (R270, R425, R635, R817, and R1000) and one never-rationed group (NR0) for whom complete data for all variables were available (n = 2960). Preferences for sweetness were represented by a latent variable, which was derived from individuals’ liking score for various sweet items such as biscuits, cake, cake icing, croissants, dark chocolate, cheesecake, jam, marzipan, milk chocolate, and other sweet foods. The latent variable for sugar consumption was based on the daily intake of free sugars and total sugars. The latent variable for brain MRI traits was derived from combining grey matter volume (GMV) measurements of specific brain regions that showed significant differences between the never-rationed group (NR0) and the five rationed group in post hoc tests. Confirmatory factor analysis was applied to estimate these latent variables. The SEM included several specified regressions and covariances. Regression equations in SEM help to model direct relationships between independent and dependent variables. The regressions were as follows: anxiety/depression was predicted by the rationing group; anxiety/depression was predicted by inflammation level, as measured by C-reactive Protein (CRP); anxiety/depression was predicted by brain MRI traits; sugar intake was predicted based on sweet-foods liking; and inflammation was predicted by sugar intake. Additionally, covariances between these variables were specified to account for potential correlations not directly modelled by the regressions. Covariances included are between sweet liking and the rationed group, sugar intake and the rationed group, sweet liking and Brain MRI traits, and Brain MRI traits and inflammation level. Model fitness was evaluated using the Root Mean Square Error of Approximation (RMSEA) and the Standardised Root Mean Square Residual (SRMR). The analyses were performed using the ‘lavaan 0.6–19’ package in R [30].

## RESULTS

Among the eligible 46,448 UK Biobank participants that met the criteria for this study, 37,229 participants were grouped into “rationed groups” and 9,219 were grouped into the never-rationed groups. The average follow-up time was 15.72 years, during which 2,776 (5.98%) participants experienced anxiety events and 3,147 (6.78%) experienced depression events. The characteristics of each group are presented in Supplementary Table 1.

### Early-life sugar rationing relates weakly to adult sweet food preferences but not sugar consumption

The correlation between the liking of sweet foods, sugar intake, and imposed sugar rationing status during gestation and infancy was examined to determine if early-life sugar rationing influences later-life sugar preferences. The overall pattern of sugar consumption (in adulthood) was consistent across rationing groups. The mean daily intake of free sugar, ranged from 58.1 to 59.2 g/day, while total sugar intake ranged from 122.4 to 123.5 g/day (Fig. 1a–b). No statistically significant differences in sugar consumption were observed between groups. Detailed distributions of free and total sugar intake stratified by mental health conditions are provided in Supplementary Table 2. Average liking scores for sweet foods were also similar across groups, with values between 5.82 and 6.04 on a 9-point Likert scale (Fig. 1c). The only significant difference was detected between the R1000 group (mean = 5.82) and the NR455 group (mean = 6.04; p = 0.0042). No other group comparisons reached statistical significance.

Analyses indicated a clear positive association between sweet liking and sugar intake (Fig 1d). In contrast, rationing status showed no meaningful association with either sugar intake or sweet food preferences, with coefficients close to zero (Fig. 1e). For example, correlations with free sugar and total sugar intake ranged between −0.005 and +0.008, and associations with specific sweet foods (e.g., biscuit, cake, chocolate) were similarly weak. These findings indicate that rationing status was not strongly associated with sugar intake or liking for individual sweet foods later in life, and no meaningful linear relationship was observed. However, inspection of the patterns suggests that the tendency of liking sweet-tasting foods and level of sugar consumption may differ between groups ever exposed to rationing, particularly R1000 and R817 groups, and those who were in never-rationed groups (NR0 and NR455). Although these differences did not reach statistical significance, they may nonetheless indicate subtle ways in which early-life rationing exposure shapes later dietary preferences.

### Association between early-life exposure to sugar and risk for anxiety and depression

The absolute risk of anxiety and depression across different groups were calculated to provide a clear measure of the likelihood of these conditions occurring within each group. The absolute risk of anxiety varied modestly across groups, ranging from 5.45% to 6.29%. The lowest risk was observed in the R817 group (5.45%), and the R1000 group (5.79%). The highest anxiety risk was found in the NR0 group (6.29%), similar to NR455 group (6,28%). Depression risks were consistently higher than anxiety, spanning 6.00% to 7.23%. The R817 group had the lowest risk (6.00%), whereas the R1000 group exhibited a risk of 6.69%. The greatest depression risk was recorded in the NR0 group (7.23%) (Supplementary Table 3).

To investigate those differences in anxiety risk across rationing status groupings, Weibull accelerated failure time (AFT) models were converted to their proportional hazards representation, with NR455 designated as the reference group (Figure 3). In the fully adjusted Weibull hazard models (Model 3), significant differences in mental health outcomes were observed across most of the rationing groups. For anxiety, early rationing exposure was consistently protective: R1000 (HR = 0.67, 95% CI: 0.53–0.84, adj p = 0.0015), R817 (HR = 0.70, 95% CI: 0.52–0.94, adj p = 0.045), and R635 (HR = 0.73, 95% CI: 0.58–0.91, adj p = 0.020) all showed significantly reduced hazards of onset. Later rationing groups (R425, R270) and NR0 did not differ from NR455. A similar pattern was evident for depression: R1000 (HR = 0.71, 95% CI: 0.57–0.87, adj p = 0.0026), R817 (HR = 0.73, 95% CI: 0.56–0.96, adj p = 0.036), and R635 (HR = 0.76, 95% CI: 0.62–0.95, adj p = 0.025) were associated with lower hazards, while R425, R270, and NR0 showed no significant differences. The examined negative control diseases—type 1 diabetes mellitus (T1DM) an
d PTSD—showed no significant associations with sugar rationing duration, suggesting that the protective effects of early-life sugar rationing may be specific to mental health outcomes and do not extend to other conditions examined here (Supplementary Figure 1). Collectively, these findings indicate that prolonged early-life rationing (R1000, R817, R635) was linked to delayed onset of both anxiety and depression, whereas shorter exposure or no rationing conferred no measurable benefit.

**Figure 2.**
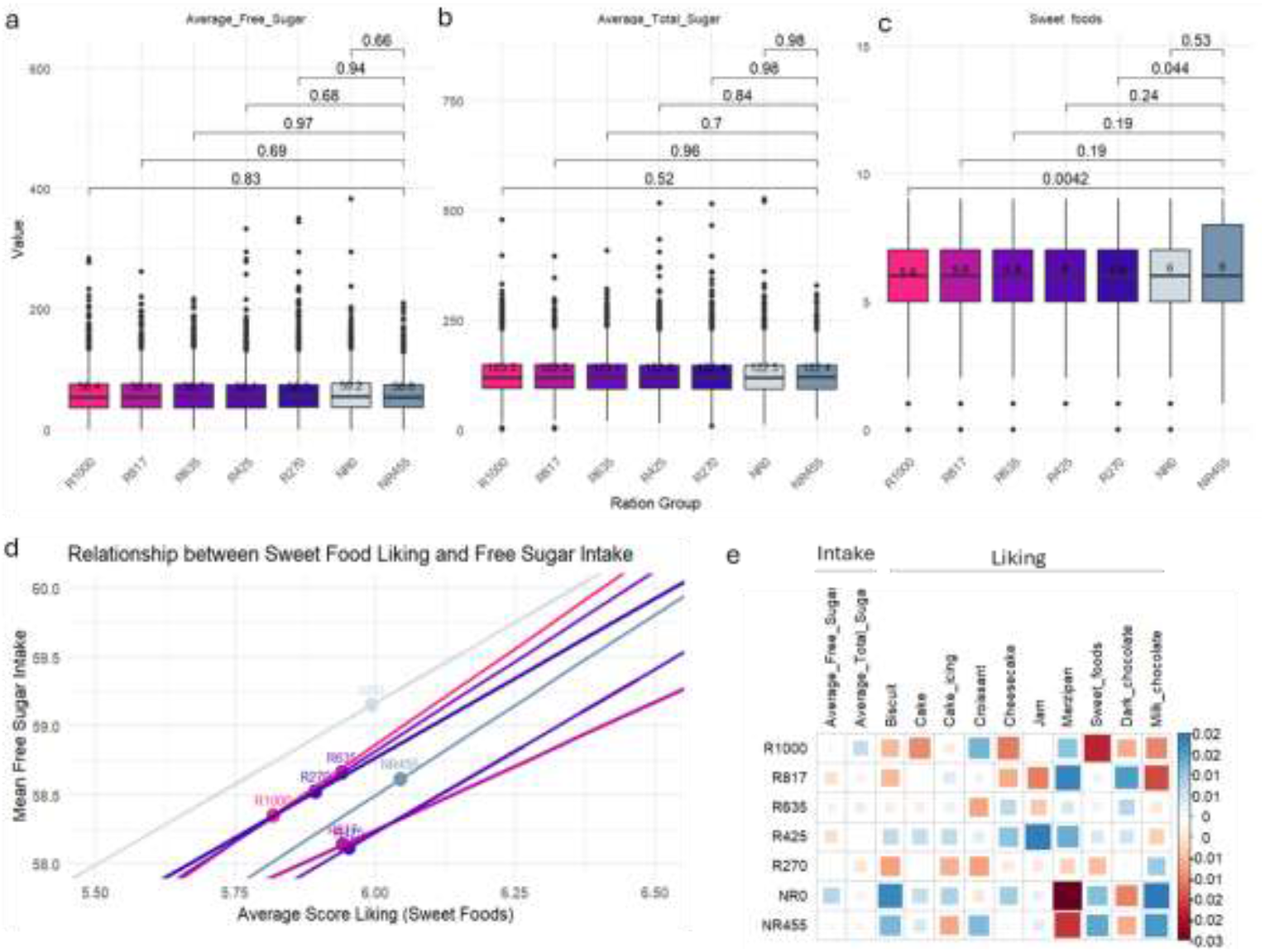
Relationship between sugar rationing status, sweet liking, and actual sugar intake. Distribution of (a) free sugar intake, (b) total sugar intake, and (a) sweet food liking scores across the seven rationing groups. (d) Relationship between average sweet food liking score and average free sugar intake. (e) Correlation plot of sugar intakes and liking for sweet foods across the seven rationing groups. Square size and colour correspond to the correlation value.

**Figure 3.**
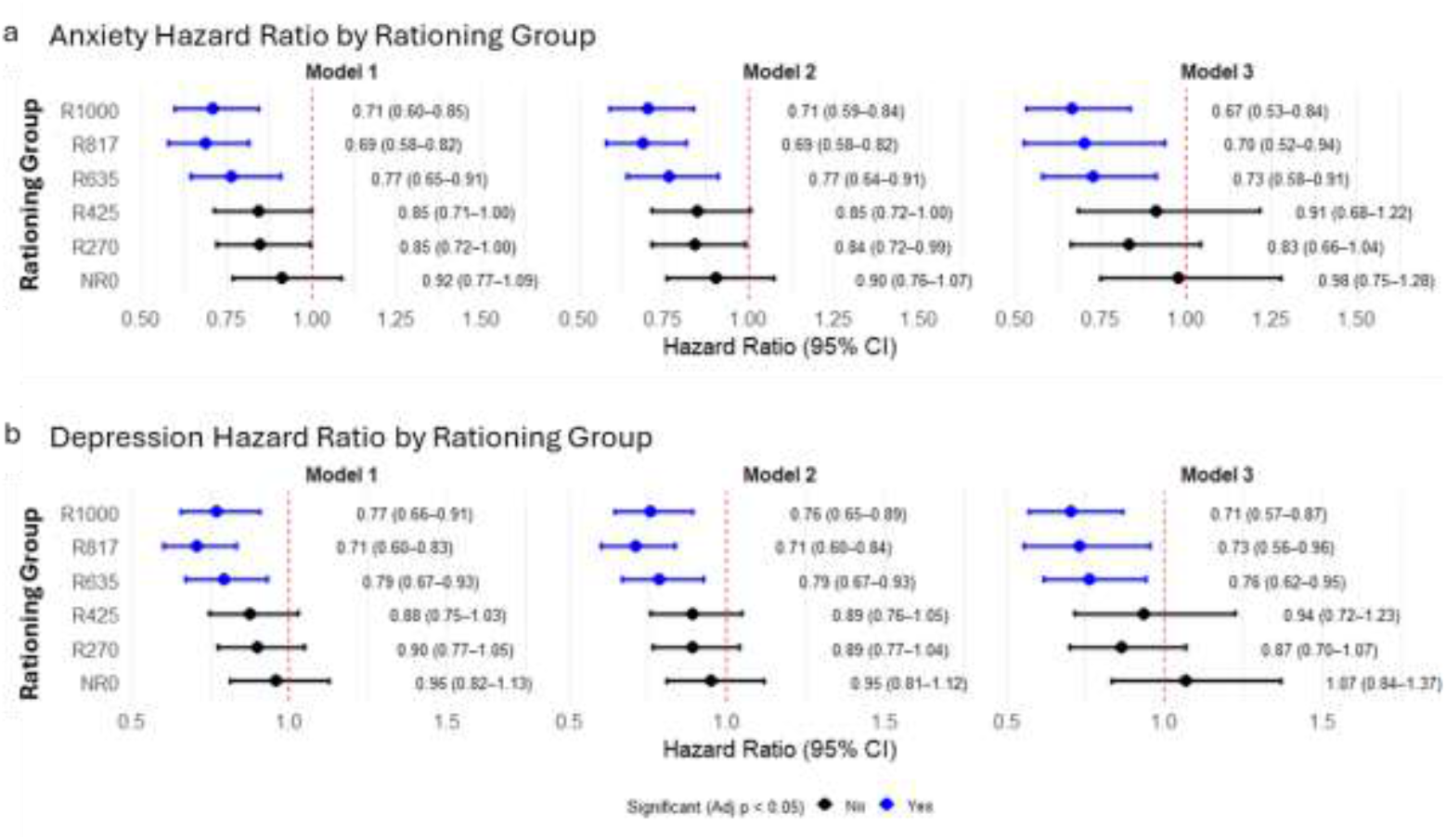
Hazard Ratio for anxiety and depression. Model 1 = unadjusted model; Model 2 = adjusted with sociodemographic factor (sex, BMI, Townsend quintiles) and season (month of birth, survey year); Model 3 = adjusted with sociodemographic, season, health factor (family history of mental health, inflammation, breastfed as child), and lifestyle (smoking status, household size, alcohol intake, sleep duration). P-value adjusted using Benjamini-Hochberg correction.

Several robustness checks were performed. In the placebo comparison group born in Ireland (NRIR), no significant associations were observed with either anxiety or depression, indicating that the protective effects identified in the UK rationing cohorts are not explained by birth timing alone but appear specific to the context of early-life rationing exposure (Supplementary Table 4). Sensitivity analyses addressing missing data yielded results consistent with the main findings, with protective associations evident for R1000, R817, and R635, and no significant associations for later groups or NR0 (Supplementary Table 5). Interaction analyses further showed that protective effects were stronger in women, with significant gender modification for R1000, R425, and NR0. Finally, additional adjustment for later-life free sugar and total sugar consumption revealed that protective associations for anxiety remained significant (notably R1000, HR = 0.63, 95% CI: 0.44–0.89, p = 0.011), whereas associations for depression were attenuated and no longer statistically significant, suggesting that adult dietary behaviour may partly account for the observed effects on depression (Supplementary Table 6). Together, these robustness checks confirm that the protective associations of early-life rationing exposure are not driven by birth timing, missing data patterns, or gender imbalance. The statistical significance attenuation observed for depression after sugar adjustment implies that dietary behaviour may play a role in shaping later-life depression risk, while the persistence of significant protection for anxiety supports the interpretation that early-life nutritional exposures exert long-term effects independent of adult sugar intake.

Differences in the median age of onset for anxiety and depression highlight the potential long-term impact of early-life rationing on mental health (Fig. 4). Median age at diagnosis varied across rationing groups for both anxiety and depression. For anxiety, individuals in the R1000 and R425 groups had the highest median onset age (61.3 years), followed by R817 (62.1), R635 and R270 (60.1), NR0 (59.5), and NR455 (55.6). For depression, median onset ages were consistently lower, with R1000 (59.1), R817 (58.4), R635 (57.5), R270 (57.9), R425 (56.8), NR0 (56.9), and NR455 (54.6). Across all groups, depression tended to be diagnosed earlier than anxiety, and those exposed to early-life rationing (R1000, R817) showed later onset compared to never-rationed groups (NR455, NR0), suggesting a possible delay in mental health vulnerability among rationed cohorts.

**Fig 4.**
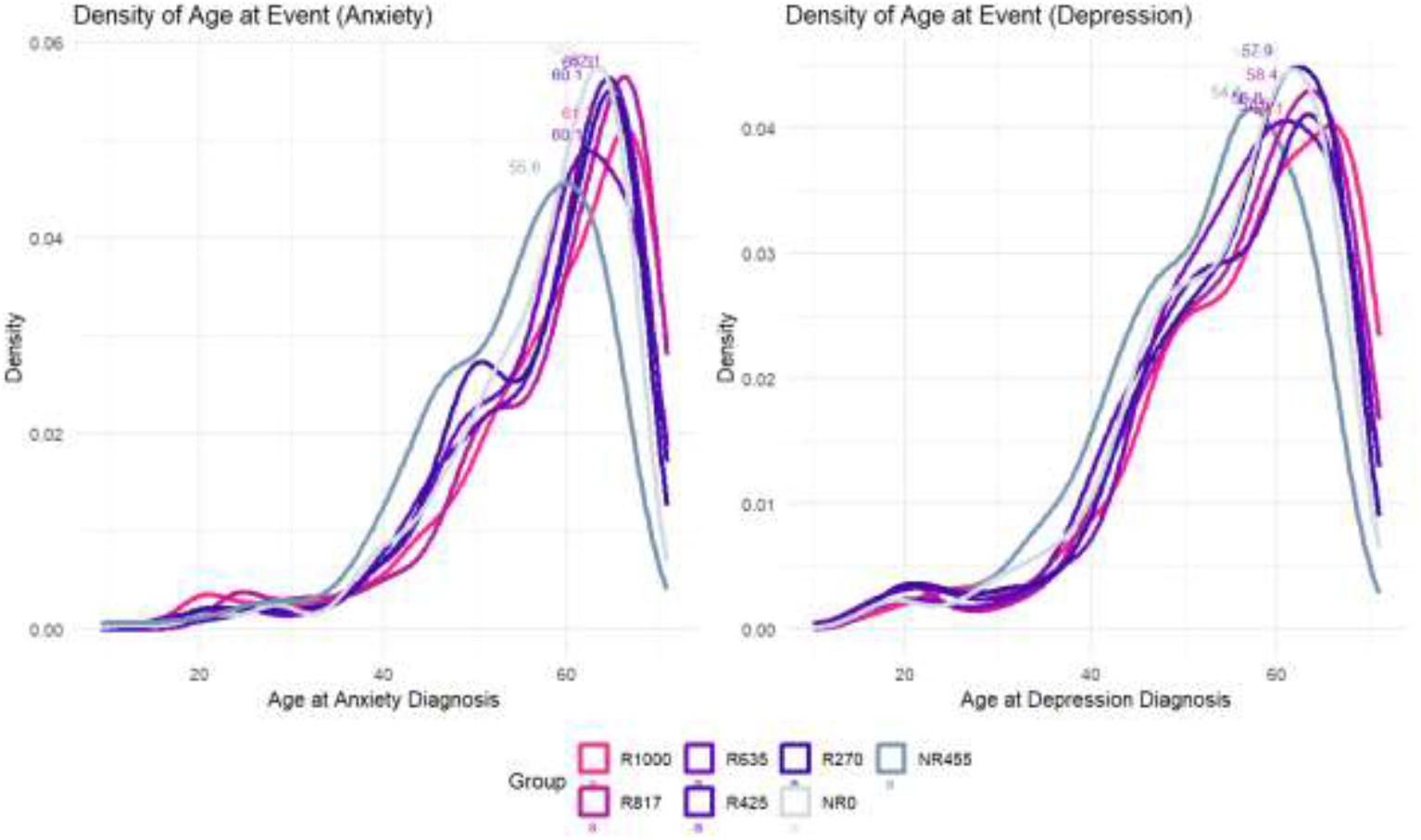
Density of age at event for (a) Anxiety and (b) Depression across Rationing Status. The x-axis represents age at diagnosis in years, while the y-axis shows the probability density, indicating the relative concentration of diagnoses at different ages. Each curve corresponds to a rationed group, with peaks marking the ages where diagnoses were most common.

### Differences in neuroimaging phenotypes across rationing status groups

We examined UK Biobank data from brain imaging in tandem with the rationing data. In one-way ANCOVA analyses, 80 of the 139 brain regions of grey matter volume were significantly different among the different rationing status groups, after applying FDR correction for multiple comparisons (adjusted P value <0.05). The assumptions of the equality of variances were satisfied for conducting one-way ANCOVAs (P > 0.05) (Supplementary Table 7). Post hoc analysis further revealed significant differences in 11 out of the 80 brain regions between most of the rationing groups and NR0 after FDR correction (adjusted P value <0.05). These regions included the brain stem, occipital fusiform gyrus and different regions of cerebellum (Fig. 5). These 11 regions were next used for creating a latent Brain MRI Trait for SEM analysis.

**Fig 5.**
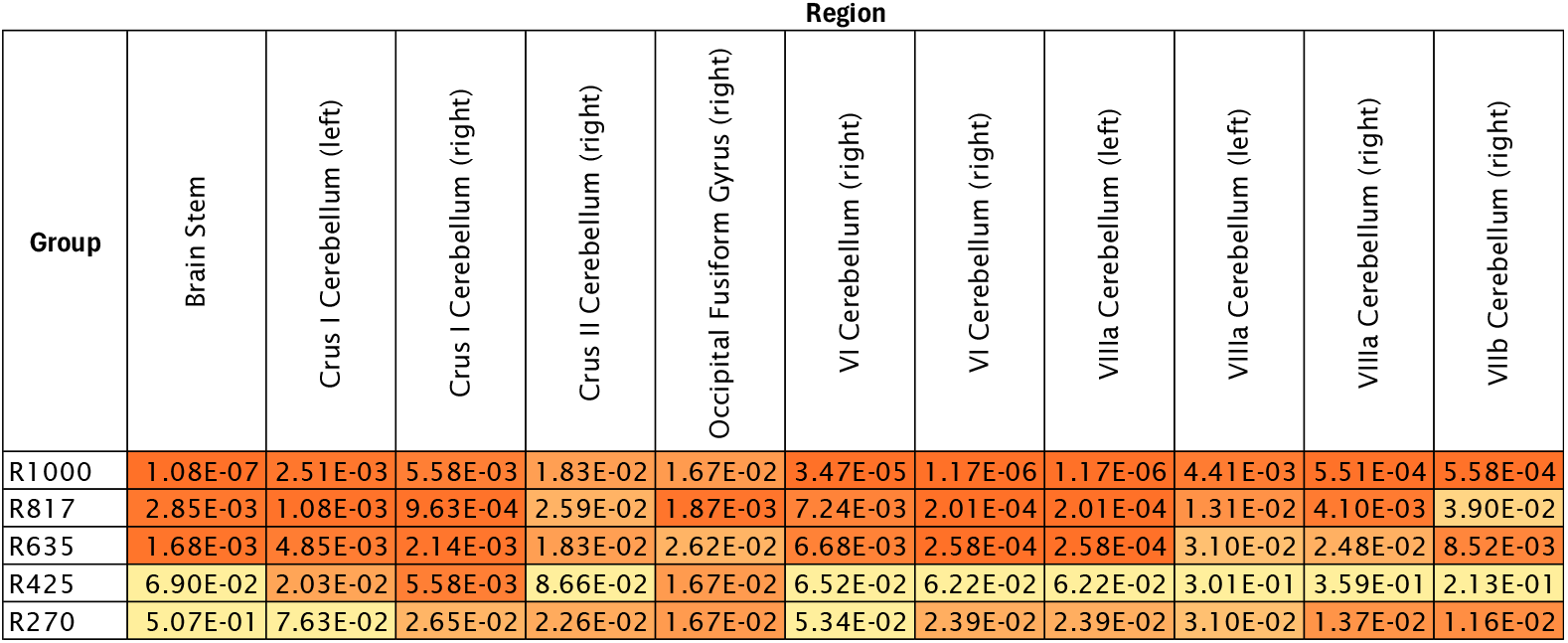
Heatmap showing significant differences in brain regions between rationing groups and the never-rationed group (NR0). The numbers inside each cell represent the adjusted p-values.

### Complex interplay of sugar rationing and mental health

To examine the complex relationships between sugar rationing exposure, liking of sweet foods, sugar intake, inflammation levels (as determined by CRP measurement), brain MRI traits, and mental health outcomes, we performed a SEM analysis. The model shows a good fit to the data, with an RMSEA of 0.057 and SRMR of 0.059 (Fig. 6). The estimates for the latent variables (sweet foods liking, sugar intake, and brain MRI traits) show significant factor loadings, indicating that the observed variables are good indicators of the latent constructs (Supplementary Table 8 and Supplementary Figure 2). The relationship between sweet taste liking and sugar intake is suggestive of a positive association between preference and intake (φ = 0.18, p < 0.001). Additionally, the covariance between sweet taste liking and brain MRI traits is significant (γ= 0.06, p = 0.003), indicating a modest but meaningful link between taste preference and structural brain traits.

**Figure 6.**
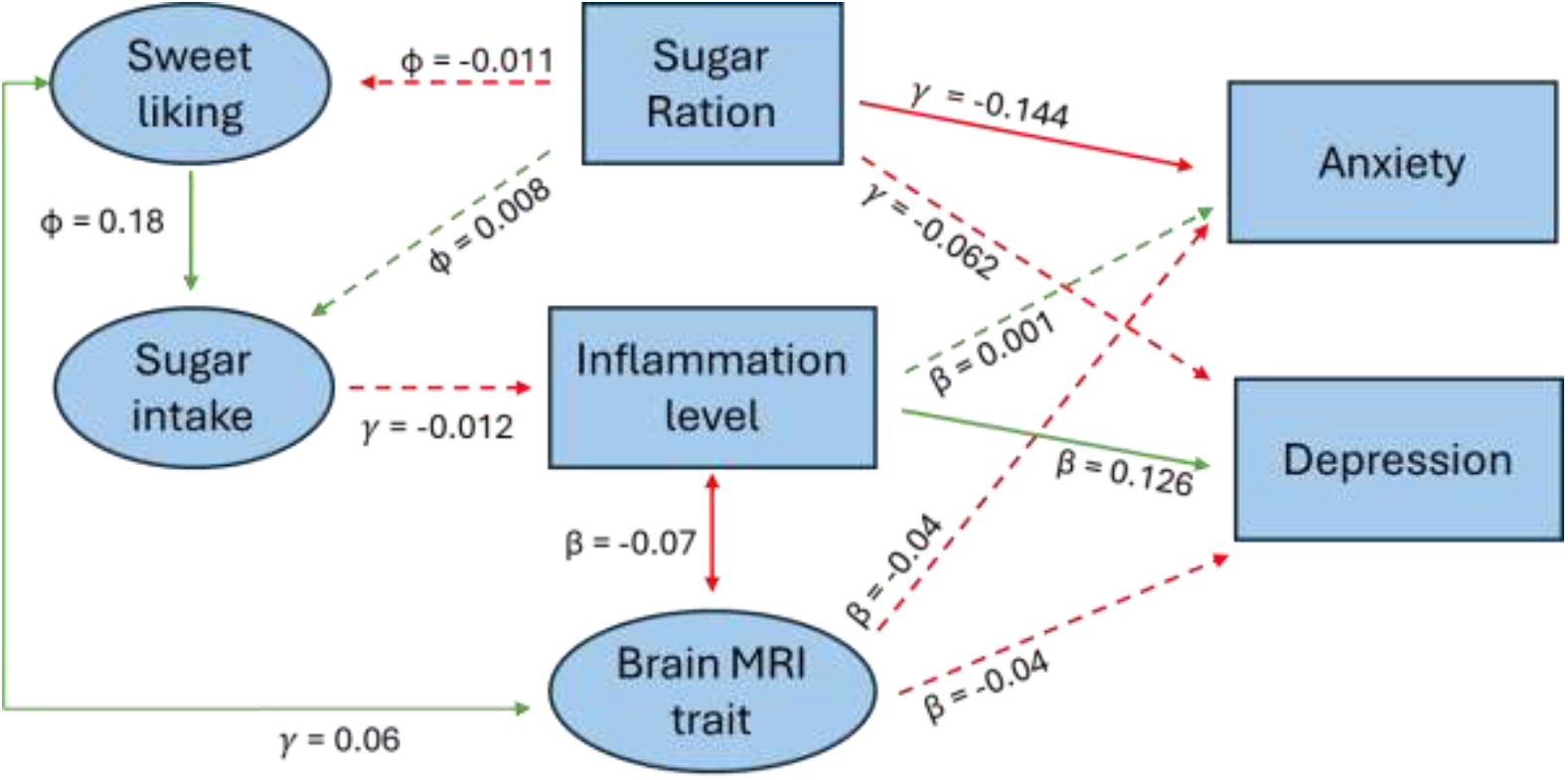
Directional associations among food preference, sugar intake, mental health outcomes, inflammation (CRP level) and brain MRI trait. Oval shape represents a latent variable. The sugar rationing variable represents the duration of days exposed to sugar rationing. The inflammation level is indicated by the CRP measurement. Anxiety and Depression is captured as a binary outcome, indicating either presence or absence. The beta (β) coefficients represent paths between endogenous variables. The gamma (*γ*) coefficients represent paths from exogenous variables to endogenous variables. The phi (φ) value represents variance or covariance of the exogenous variable. The magnitude of the coefficient indicates the strength of the relationship, while the sign (positive or negative) indicates the direction. Red arrows represent negative correlation; Green arrows represent positive correlation. Dashed Arrow: Non-significant or weaker relationships. Solid Arrow: Significant or strong relationships. One-Direction Arrow: Causal relationship from one variable to another. Two-Direction Arrow: Correlation or bidirectional relationship.

The direct effect of rationing status on anxiety is significant and negative (*γ*= -0.144, p = 0.012), suggesting that shorter durations of early-life sugar rationing are associated with a higher likelihood of anxiety occurrence in adulthood. In contrast, the direct effect of rationing status on depression is not statistically significant (*γ*= -0.062, p = 0.241). Furthermore, the indirect effects through inflammation and brain MRI traits are not significant. The regression coefficient for the direct effect of rationing status on depression is -0.062 (p = 0.241), indicating no strong evidence for a direct causal relationship between rationing status and depression. Furthermore, indirect pathways through inflammation and brain MRI traits were not statistically significant for either anxiety or depression. Specifically, the hypothesized pathway from sugar intake to depression via inflammation was unsupported, as neither the path from sugar intake to inflammation nor from inflammation to depression reached significance. Similarly, the indirect pathway from rationing status to depression via brain MRI traits was not supported, given the non-significant association between brain MRI traits and depression. Nonetheless grey matter volume was significantly different especially between the prolonged rationing groups and the never rationed groups.among the rationed status groups,

## DISCUSSION

This study aimed to explore the long-term effects of early-life sugar rationing (and thus, inversely, exposure to sugar) on later-life sugar preferences, brain structure, and mental health outcomes. This study provides compelling evidence that early-life sugar rationing has lasting effects on preferences for sweet foods, mental health outcomes, and brain structure, highlighting the critical role of early dietary experiences in shaping long-term health and behaviour. Participants exposed to the longest duration of rationing (R1000) reported lower preference for sweet foods and showed significantly reduced hazards of anxiety in adulthood, independent of later-life dietary behaviours. A protective association with depression was also evident, though this was attenuated in sensitivity analyses that accounted for later-life sugar intake, indicating that subsequent diet may partly mediate this relationship.

Although sugar was not the only food rationed during this period—other staples such as cheese, butter, cereal, and meat were also restricted—the end of sugar rationing occurred earlier than that of most other foods. Further, once sugar was reintroduced, this had significantly higher uptake, as compared to uptake of other foods when those became more freely available[12]. Nevertheless, to better isolate the effects of sugar rationing from the broader context of post-war food restrictions, we selected NR455 (conceived one year after all food rationing ended) as the reference group rather than NR0 (conceived immediately after sugar rationing ended). The NR0 group, while unexposed to sugar rationing, was born during a period when other foods remained rationed, which may have introduced residual nutritional or socioeconomic influences. Hazard ratios for NR0 relative to NR455 were close to null for both anxiety and depression, indicating that residual rationing of other foods did not materially affect mental health outcomes. This supports the rationale for using NR455 as the cleaner baseline, ensuring that the protective associations observed in the rationed groups (R1000, R817, R635) can be more confidently attributed to sugar rationing rather than to broader wartime food scarcity. Against this backdrop, the protective associations observed in the R1000 group highlight sugar’s distinct role in shaping dietary preferences and mental health outcomes. While these patterns align with our hypothesis linking early-life sugar exposure to long-term mental health, residual confounding from unmeasured socioeconomic, environmental, or diagnostic factors cannot be fully excluded. Findings should therefore be interpreted with appropriate caution, but they underscore the potential importance of sugar exposure specifically in influencing later-life mental health risk.

Building on this rationale, it is important to situate our findings within the broader historical and contemporary context of sugar consumption and its established links to mental health. Historical data indicate a consistent increase in per capita sugar consumption in the United Kingdom since the cessation of food rationing until 1978, with a notable acceleration following the introduction of artificial sweeteners widely used in soft drinks, pastries, and other processed foods [31]. Children, in particular, have historically been disproportionately exposed to high levels of free sugars and ultra-processed foods—contributing to elevated health risks that peaked in the early 2000s. Although intake of sugar-sweetened beverages and added sugars began to decline thereafter in response to growing public health concerns [32,33], current levels of free sugar intake among UK adults remain above recommended thresholds. According to the National Diet and Nutrition Survey (2019–2023), current free sugar intake across all age groups in the UK continues to exceed government recommendations that no more than 5% of total energy should come from free sugars; only 9% of children and 19% of adults meet this target, with mean intakes in most age groups—particularly among adolescents and working-age adults—hovering around double the recommended level [34]. These contemporary data demonstrate that despite decades of public health efforts, the legacy of historically high sugar consumption persists, with current intakes still far above recommended limits. This continuity underscores the relevance of examining early-life sugar rationing: by contrasting a period of enforced restriction with a period reflective of today’s environment of excess, our findings highlight how nutritional exposures during critical developmental windows may shape resilience or vulnerability to mental health outcomes across generations.

To test whether effects could be explained simply by birth timing, we included a comparison group born in Ireland (NRIR), comprising adults born during the same period as R1000 and R817 but where sugar was not rationed at that time. Although Ireland also maintained rationing into the early 1950s, its policies differed from those in the UK. No significant associations were observed with either anxiety or depression in this group, indicating that the protective effects of sugar rationing identified in the UK cohorts are not explained by timing of birth alone but appear specific to the UK rationing context. Sensitivity analyses also showed that adjusting for later-life sugar intake reduced the effect size of rationing, suggesting that part of the association may be driven by subsequent dietary exposure. Across most rationing groups, individuals with anxiety consumed more sugar than those without, though this pattern was not consistent in all groups. These findings align with prior work linking sugar and mental health [7,8,35], with evidence that the relationship is bidirectional [36], while also underscoring the complexity of these associations. As the dietary analysis was limited to a subset differing in age, BMI, deprivation, and other characteristics, selection bias remains possible. Taken together, these observations highlight the interplay between early-life exposures, later-life diet, and broader contextual factors in shaping mental health outcomes.

To explore possible causal explanations for how a longer duration of sugar rationing contributes to the protective effect, we developed a model assessing associations between mental health outcomes (anxiety and depression), inflammation, brain morphology, and exposure to sugar rationing duration. We found a significant positive association between inflammation levels, measured by C-reactive protein (CRP), and depression, but not with anxiety. This aligns with previous studies suggesting that inflammatory processes contribute more consistently to depressive symptoms than to anxiety [37]. However, our analysis was limited to CRP and did not include other inflammatory markers such as interleukin-6 (IL-6), which several studies have found more reliably associated with depression [38,39]. We therefore attempted to establish an indirect pathway linking sugar rationing to depression through sugar intake and inflammation, but no significant association emerged between rationing duration and sugar intake. Thus, while inflammation appears to play a role in depression, it does not explain the protective effects of sugar rationing, prompting us to examine neurodevelopmental mechanisms through brain morphology.

The robust association between sugar rationing and reduced anxiety risk underscores the importance of early nutritional environments in shaping neurodevelopmental trajectories. Critical periods in the first 1000 days of life represent windows of heightened plasticity, during which nutrient availability can calibrate neuronal growth, synaptic connectivity, and long-term stress regulation [40–42]. In an important and relevant finding worthy of further investigation we show that sugar rationing during this sensitive window may have induced lasting changes in brain structure and function, which in turn could shape both dietary preferences and protection from mental health conditions. A diet high in sugar affects neurotransmitter metabolism in mice models [43]. Indeed, differences in grey matter volume observed in relation to rationing exposure align with the notion that early nutritional deprivation can alter dendritic complexity and synaptic efficacy, with consequences persisting into adulthood. Prior evidence further supports this pathway, showing that food preferences, brain morphology, and mental health are interconnected [44]. In our SEM analyses, sweet liking was associated with brain MRI traits, but sugar rationing itself did not directly predict sweet preference, suggesting that altered taste preference is unlikely to fully explain the observed associations. Taken together, these results imply that mechanisms beyond dietary preference—potentially involving neurodevelopmental programming during critical periods—may underlie the protective effects of early-life sugar exposure on mental health outcomes.

Our findings need to be considered in the context of study limitations. Although the UK Biobank collects genomic information, DNA methylation data are not yet available, thus examining whether epigenetic changes mediate the protective effects of sugar rationing was not possible. While we could not confirm such mechanisms in our study, recent findings by van den Berg et al. (2025) suggest that prenatal sugar exposure can interact with genetic predisposition to shape later-life outcomes, including education, BMI, and sugar consumption [13]. Secondly, while SEM enables the testing of theoretically informed causal models, the observational nature of the data limits definitive causal inference, and residual confounding from unmeasured or inadequately controlled variables cannot be ruled out. Lastly, because our analyses focus on participants born in the UK during the rationing period, the generalisability of these findings to other populations may be limited. Although Ireland and other countries also implemented sugar rationing, the timing, duration, and policy context differed, meaning the protective effects observed here may be specific to the UK experience.

This study demonstrates that limiting sugar exposure during the first 1000 days of life exerts enduring effects on sweet food preferences and psychiatric outcomes. Early-life rationing was associated with reduced sweet liking and a lower risk of anxiety in adulthood, whereas depression appeared more closely linked to inflammatory pathways. The absence of a direct association between rationing and sweet preference suggests that protective effects are likely mediated through neurodevelopmental programming during critical developmental periods. Despite increased sugar consumption following the end of rationing, our findings underscore the long-term influence of early nutritional environments on mental health and behaviour. Future research should incorporate epigenetic and gene–environment analyses, expanded inflammatory profiling from forthcoming UK Biobank proteomic data, and neuroimaging approaches to elucidate the mechanisms by which early-life nutrition shapes psychiatric resilience across populations.

## Acknowledgements

The study utilised data from UK Biobank, see http://www.ukbiobank.ac.uk. We thank UK Biobank volunteers for their involvement.

## Contributor and guarantor information

HFN contributed to the data analysis, interpretation of the results, and drafting the article. ADW contributed to the interpretation of the results and critically revised the manuscript for important intellectual content. NG contributed to the conception and design of the study, interpretation of the results, and critically revised the manuscript for important intellectual content. The corresponding author attests that all listed authors meet authorship criteria and that no others meeting the criteria have been omitted. NG is guarantor of the paper.

## Ethical approval

This study uses UK Biobank data under existing ethical approval (REC 16/NW/0274), overseen by the UK Biobank Ethics Advisory Committee, and has been confirmed by the University of Surrey Research Integrity & Governance Office (FHMS 22‐23 273 EGA) as not requiring additional ethical approval.

## Consent to participate

Written informed consent was obtained for all UK Biobank study participants.

## Data availability

Our study utilised the UK Biobank under research application number 83988. The raw data used in this study are not publicly available due to restrictions imposed by the UK Biobank to protect participant privacy. Researchers can apply for access to the UK Biobank data through the official application process at https://www.ukbiobank.ac.uk. The analysis code used for this study was tailored to the UK Biobank data and is no use as a standalone without access to the UK Biobank (https://github.com/hanavratilova/sr_ukb).

