## Supplementary figures and images for "Dietary sugar exposure in early life and risk of adult mental health disorders: UK Biobank cohort study"

### Supplementary Figure 1

Supplementary Figure 1. Hazard Ratio of T1DM and PTSD

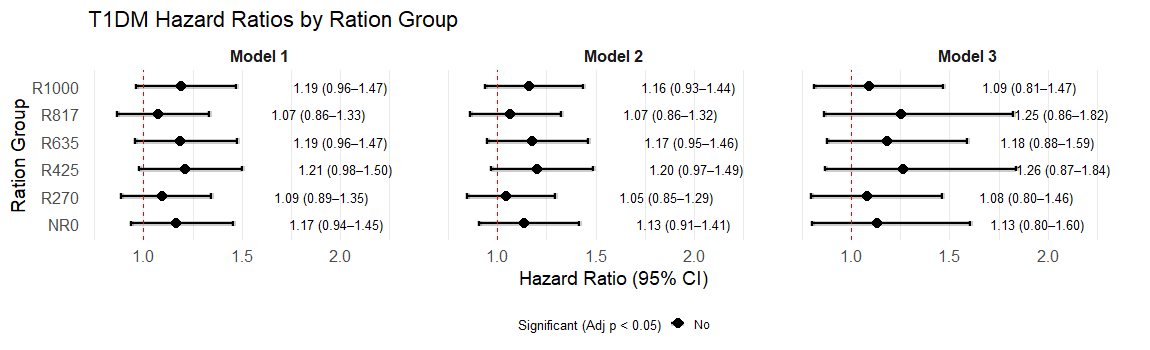

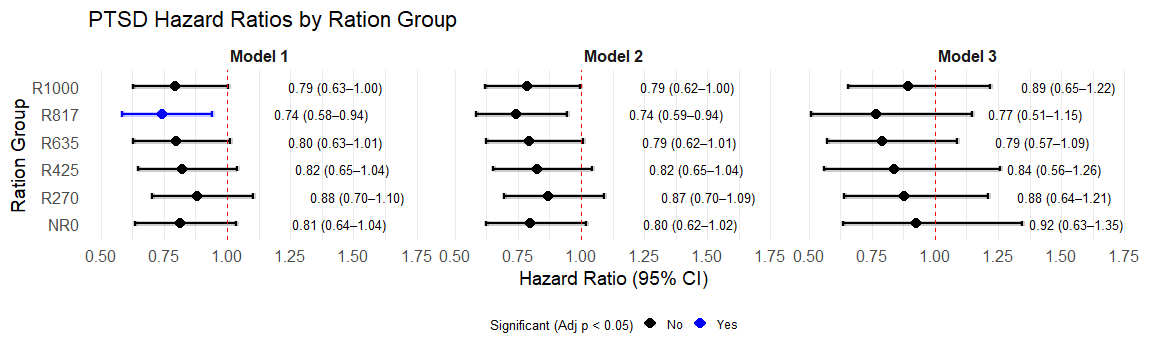

### Supplementary Figure 2

Supplementary Figure 2. SEMpath for (a) anxiety and (b) depression

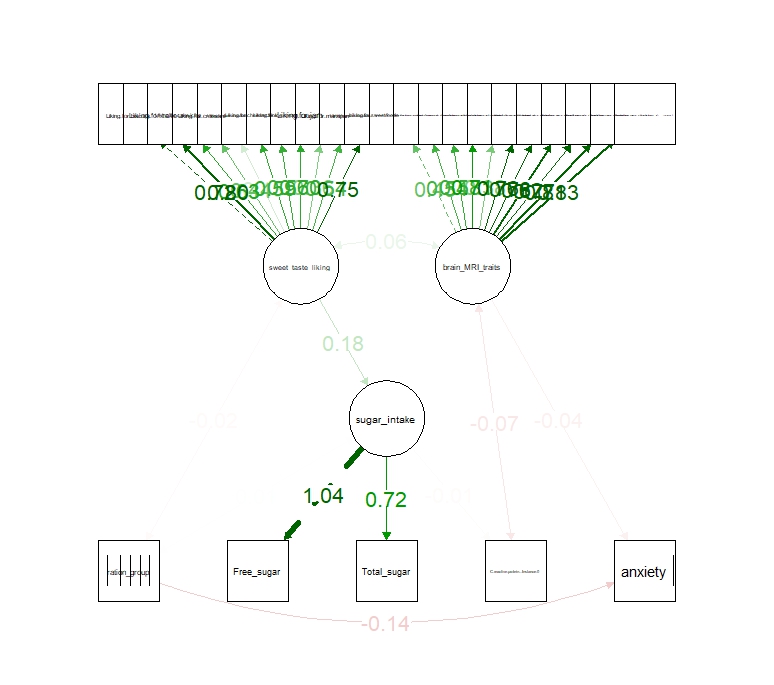

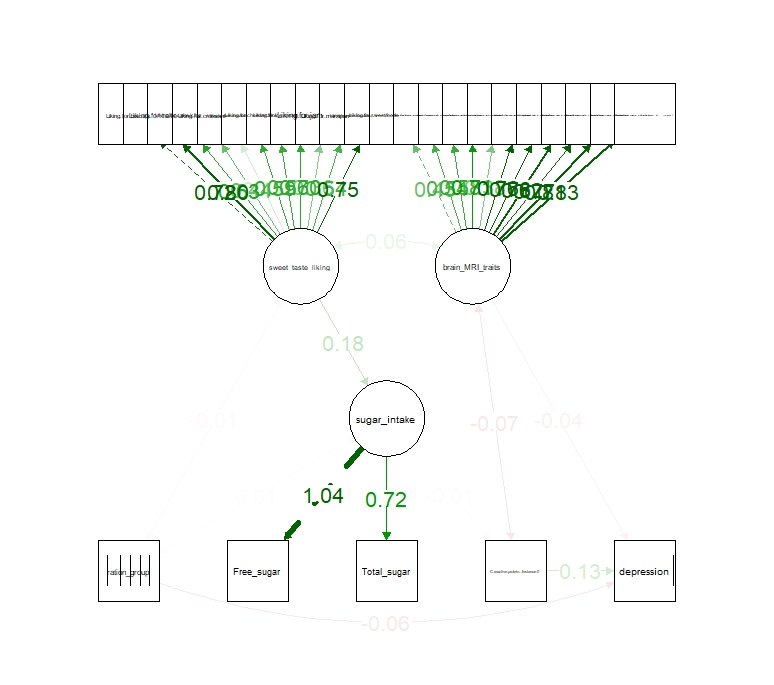
