## Supplementary Table 1 for "Dietary sugar exposure in early life and risk of adult mental health disorders: UK Biobank cohort study"

Supplementary Table 1. Baseline characteristics

| **Characteristic** | **Overall** | **NR455** | **R1000** | **R817** | **R635** | **R425** | **R270** | **NR0** | **p-value***^2^* |
| --- | --- | --- | --- | --- | --- | --- | --- | --- | --- |
|  | N = 46,448*^1^* | N = 3,102*^1^* | N = 6,856*^1^* | N = 7,096*^1^* | N = 6,622*^1^* | N = 6,984*^1^* | N = 9,671*^1^* | N = 6,117*^1^* |  |
| Sex |  |  |  |  |  |  |  |  | 0.7 |
| Female | 25,796 (56%) | 1,761 (57%) | 3,829 (56%) | 3,957 (56%) | 3,652 (55%) | 3,847 (55%) | 5,342 (55%) | 3,408 (56%) |  |
| Male | 20,652 (44%) | 1,341 (43%) | 3,027 (44%) | 3,139 (44%) | 2,970 (45%) | 3,137 (45%) | 4,329 (45%) | 2,709 (44%) |  |
| townsend_quintiles |  |  |  |  |  |  |  |  | 0.6 |
| Q1 | 16,871 (36%) | 1,115 (36%) | 2,472 (36%) | 2,557 (36%) | 2,421 (37%) | 2,576 (37%) | 3,528 (37%) | 2,202 (36%) |  |
| Q2 | 10,393 (22%) | 684 (22%) | 1,522 (22%) | 1,588 (22%) | 1,497 (23%) | 1,572 (23%) | 2,155 (22%) | 1,375 (22%) |  |
| Q3 | 7,694 (17%) | 513 (17%) | 1,101 (16%) | 1,213 (17%) | 1,080 (16%) | 1,192 (17%) | 1,603 (17%) | 992 (16%) |  |
| Q4 | 6,575 (14%) | 455 (15%) | 989 (14%) | 1,029 (15%) | 911 (14%) | 966 (14%) | 1,360 (14%) | 865 (14%) |  |
| Q5 | 4,853 (10%) | 333 (11%) | 761 (11%) | 706 (10.0%) | 703 (11%) | 666 (9.6%) | 1,006 (10%) | 678 (11%) |  |
| family_history | 4,209 (9.1%) | 269 (8.7%) | 607 (8.9%) | 634 (8.9%) | 576 (8.7%) | 687 (9.8%) | 891 (9.2%) | 545 (8.9%) | 0.3 |
| breastfed | 27,340 (74%) | 1,818 (72%) | 4,068 (75%) | 4,297 (77%) | 3,882 (73%) | 4,061 (74%) | 5,670 (73%) | 3,544 (72%) | <0.001 |
| bmi_class |  |  |  |  |  |  |  |  | 0.024 |
| Normal | 15,349 (33%) | 1,093 (35%) | 2,246 (33%) | 2,356 (33%) | 2,128 (32%) | 2,317 (33%) | 3,134 (33%) | 2,075 (34%) |  |
| Underweight | 270 (0.6%) | 28 (0.9%) | 41 (0.6%) | 35 (0.5%) | 43 (0.7%) | 34 (0.5%) | 57 (0.6%) | 32 (0.5%) |  |
| Overweight | 19,041 (41%) | 1,226 (40%) | 2,792 (41%) | 2,973 (42%) | 2,788 (42%) | 2,869 (41%) | 3,963 (41%) | 2,430 (40%) |  |
| Obesity | 11,600 (25%) | 745 (24%) | 1,756 (26%) | 1,708 (24%) | 1,626 (25%) | 1,743 (25%) | 2,470 (26%) | 1,552 (25%) |  |
| alcohol_intake |  |  |  |  |  |  |  |  | <0.001 |
| Never | 2,588 (5.6%) | 177 (5.7%) | 430 (6.3%) | 380 (5.4%) | 387 (5.8%) | 395 (5.7%) | 487 (5.0%) | 332 (5.4%) |  |
| Special occasions only | 4,490 (9.7%) | 266 (8.6%) | 734 (11%) | 681 (9.6%) | 668 (10%) | 649 (9.3%) | 933 (9.7%) | 559 (9.1%) |  |
| One to three times a month | 5,032 (11%) | 331 (11%) | 735 (11%) | 752 (11%) | 729 (11%) | 749 (11%) | 1,050 (11%) | 686 (11%) |  |
| Once or twice a week | 12,654 (27%) | 904 (29%) | 1,804 (26%) | 1,920 (27%) | 1,808 (27%) | 1,909 (27%) | 2,626 (27%) | 1,683 (28%) |  |
| Three or four times a week | 11,880 (26%) | 819 (26%) | 1,683 (25%) | 1,792 (25%) | 1,584 (24%) | 1,810 (26%) | 2,613 (27%) | 1,579 (26%) |  |
| Daily or almost daily | 9,779 (21%) | 604 (19%) | 1,469 (21%) | 1,565 (22%) | 1,442 (22%) | 1,470 (21%) | 1,955 (20%) | 1,274 (21%) |  |
| household_size |  |  |  |  |  |  |  |  | <0.001 |
| 1 | 7,925 (20%) | 476 (19%) | 1,227 (20%) | 1,225 (19%) | 1,167 (20%) | 1,198 (20%) | 1,663 (20%) | 969 (19%) |  |
| 2-4 | 29,766 (75%) | 1,770 (72%) | 4,623 (76%) | 4,834 (77%) | 4,257 (75%) | 4,490 (75%) | 6,019 (74%) | 3,773 (75%) |  |
| >4 | 2,041 (5.1%) | 196 (8.0%) | 250 (4.1%) | 256 (4.1%) | 276 (4.8%) | 295 (4.9%) | 447 (5.5%) | 321 (6.3%) |  |
| smoking_status |  |  |  |  |  |  |  |  | <0.001 |
| Never | 26,143 (56%) | 1,829 (59%) | 3,750 (55%) | 3,932 (56%) | 3,714 (56%) | 3,982 (57%) | 5,492 (57%) | 3,444 (56%) |  |
| Previous | 15,220 (33%) | 894 (29%) | 2,366 (35%) | 2,382 (34%) | 2,170 (33%) | 2,278 (33%) | 3,152 (33%) | 1,978 (32%) |  |
| Current | 4,947 (11%) | 368 (12%) | 718 (11%) | 759 (11%) | 722 (11%) | 706 (10%) | 999 (10%) | 675 (11%) |  |
| sleep_duration |  |  |  |  |  |  |  |  | 0.11 |
| <7 | 12,488 (27%) | 826 (27%) | 1,829 (27%) | 1,913 (27%) | 1,807 (27%) | 1,808 (26%) | 2,665 (28%) | 1,640 (27%) |  |
| 7-9 | 33,232 (72%) | 2,222 (72%) | 4,900 (72%) | 5,059 (72%) | 4,707 (71%) | 5,069 (73%) | 6,881 (71%) | 4,394 (72%) |  |
| >9 | 526 (1.1%) | 41 (1.3%) | 90 (1.3%) | 95 (1.3%) | 79 (1.2%) | 70 (1.0%) | 88 (0.9%) | 63 (1.0%) |  |
| inflammation_status |  |  |  |  |  |  |  |  | 0.9 |
| <8 | 41,000 (95%) | 2,752 (95%) | 6,063 (95%) | 6,252 (94%) | 5,851 (95%) | 6,165 (95%) | 8,499 (94%) | 5,418 (95%) |  |
| ≥8 | 2,348 (5.4%) | 156 (5.4%) | 350 (5.5%) | 364 (5.5%) | 329 (5.3%) | 338 (5.2%) | 512 (5.7%) | 299 (5.2%) |  |
| *^1^* n (%) | | | | | | | | | |
| *^2^* Pearson’s Chi-squared test | | | | | | | | | |
