## Supplementary Table 2 for "Dietary sugar exposure in early life and risk of adult mental health disorders: UK Biobank cohort study"

Supplementary table 2. Free Sugar and Total Sugar intake stratified by mental health conditions

| **Characteristic** | **R270** | **R425** | **R635** | **R817** | **R1000** | **NR0** | **NR455** | **p-value***^2^* |
| --- | --- | --- | --- | --- | --- | --- | --- | --- |
| **Free sugar (g/day)** | 58.5 (36.3, 74.5) | 58.1 (35.6, 73.9) | 58.6 (35.4, 75.6) | 58.2 (35.9, 75.4) | 58.2 (36.1, 74.6) | 59.2 (37.3, 76.0) | 58.5 (37.1, 73.6) | >0.9 |
| Anxiety |  |  |  |  |  |  |  |  |
| no | 58.5 (36.2, 74.4) | 57.8 (35.3, 73.4) | 58.4 (35.3, 75.5) | 58.4 (36.2, 75.6) | 58.0 (36.1, 74.0) | 59.0 (37.1, 75.9) | 58.6 (37.1, 73.7) |  |
| yes | 62.0 (39.3, 79.2) | 66.5 (43.4, 85.2) | 66.5 (42.5, 80.5) | 51.0 (32.2, 68.6) | 66.6 (40.1, 84.5) | 64.3 (39.4, 84.6) | 56.5 (37.6, 66.2) |  |
| Depression |  |  |  |  |  |  |  |  |
| no | 58.5 (36.3, 74.4) | 57.8 (35.6, 73.7) | 58.7 (35.4, 75.7) | 58.1 (35.9, 75.4) | 58.3 (36.3, 74.6) | 59.0 (37.3, 75.9) | 58.5 (36.9, 73.6) |  |
| yes | 59.9 (35.2, 75.2) | 64.6 (39.1, 79.5) | 57.6 (36.1, 74.1) | 59.3 (35.8, 78.0) | 54.0 (30.7, 70.9) | 64.0 (36.3, 74.6) | 61.2 (40.0, 72.0) |  |
| **Total sugar (g/day)** | 122 (92, 147) | 123 (93, 147) | 123 (92, 148) | 122 (93, 147) | 123 (94, 147) | 123 (92, 147) | 122 (93, 145) | 0.9 |
| Anxiety |  |  |  |  |  |  |  |  |
| no | 122 (91.6, 147) | 122 (93.5, 147) | 123 (92, 148) | 123 (93.1, 148) | 123 (93.6, 147) | 122 (91.7, 146) | 122 (92.6, 145) |  |
| yes | 134 (102, 158) | 130 (94.5, 156) | 132 (95.2, 160) | 115 (88.4, 135) | 125 (90.7, 154) | 137 (108, 158) | 118 (86.1, 145) |  |
| Depression |  |  |  |  |  |  |  |  |
| no | 122 (91.8, 147) | 122 (93.3, 147) | 123 (92, 148) | 123 (93.1, 147) | 124 (93.8, 148) | 122 (92.3, 146) | 122 (92.6, 145) |  |
| yes | 127 (90.0, 143) | 133 (99.5, 157) | 122 (93, 152) | 119 (88.7, 139) | 111 (86.9, 140) | 128 (91.1, 161) | 129 (91.4, 152) |  |
| *^1^* Mean (Q1, Q3) | | | | | | | | |
| *^2^* Kruskal-Wallis rank sum test | | | | | | | | |
