## Supplementary Table 3 for "Dietary sugar exposure in early life and risk of adult mental health disorders: UK Biobank cohort study"

Supplementary table 3. Absolute risk for anxiety and depression

| Group | Anxiety %absolute risk (95%CI) | | | Depression %absolute risk (95%CI) | | |
| --- | --- | --- | --- | --- | --- | --- |
|  | All | Female | Male | All | Female | Male |
| R1000 | 5.79 (5.25, 6.38) | 6.99 (6.22, 7.8) | 4.26 (3.58, 5.05) | 6.69 (6.12, 7.32) | 7.62 (6.81, 8.52) | 5.52 (4.74, 6.40) |
| R817 | 5.45 (4.94, 6.01) | 6.47 (5.73, 7.29) | 4.17 (3.51, 4.95) | 6.00 (5.47, 6.59) | 7.10 (6.33, 7.96) | 4.62 (3.92, 5.43) |
| R635 | 5.86 (5.31, 6.46) | 6.62 (5.85, 7.49) | 4.91 (4.18, 5.77) | 6.54 (5.96, 7.17) | 7.75 (6.91, 8.67) | 5.05 (4.30, 5.91) |
| R425 | 6.27 (5.72 , 6.87) | 7.61 (6.81, 8.51) | 4.62 (3.93, 5.43) | 7.04 (6.46, 7.68) | 8.06 (7.23, 8.97) | 5.80 (5.02, 6.69) |
| R270 | 6.06 (5.59, 6.56) | 7.41 (6.73, 8.16) | 4.39 (3.81, 5.05) | 7.01 (6.51, 7.54) | 16.51 (15.58, 17.49) | 11.22 (10.33, 12.17) |
| NR0 | 6.29 (5.70, 6.94) | 7.25 (6.41, 8.18) | 5.09 (4.31, 6.01) | 7.22 (6.59, 7.91) | 8.14 (7.43, 8.92) | 5.61 (4.95, 6.35) |
| NR455 | 6.29 (5.47, 7.21) | 7.21 (6.07, 8.54) | 5.07 (3.97, 6.42) | 6.99 (6.13, 7.96) | 7.95 (7.07, 8.92) | 6.31 (5.44, 7.31) |
