## Supplementary Table 4 for "Dietary sugar exposure in early life and risk of adult mental health disorders: UK Biobank cohort study"

Supplementary table 4. Robustness check (NRIR group inclusion)

|  | Anxiety | | Depression | |
| --- | --- | --- | --- | --- |
|  | HR (95% CI) | P-value | HR (95% CI) | Adjusted P-value |
| R1000 | 0.71 (0.60, 0.84) | <0.001 | 0.77 (0.65, 0.90) | 0.001 |
| R817 | 0.68 (0.79, 0.81) | < 0.001 | 0.71 (0.60, 0.83) | < 0.001 |
| R635 | 0.76 (0.64, 0.91) | 0.002 | 0.79 (0.67, 0.93) | 0.005 |
| R425 | 0.84 (0.71, 1.00) | 0.05 | 0.88 (0.75, 1.03) | 0.11 |
| R270 | 0.84 (0.72, 0.99) | 0.04 | 0.90 (0.77, 1.05) | 0.18 |
| NR0 | 0.91 (0.77, 1.08) | 0.3 | 0.96 (0.82, 1.13) | 0.62 |
| NRIR | 1.09 (0.61, 1.95) | 0.7 | 0.54 (0.22, 1.13) | 0.09 |
