## Supplementary Table 5 for "Dietary sugar exposure in early life and risk of adult mental health disorders: UK Biobank cohort study"

Supplementary table 5. Robustness check (Imputation of missing value)

|  | Anxiety | | Depression | |
| --- | --- | --- | --- | --- |
|  | HR (95% CI) | P-value | HR (95% CI) | Adjusted P-value |
| R1000 | 0.72 (0.60, 0.86) | <0.001 | 0.76 (0.64, 0.90) | 0.001 |
| R817 | 0.68 (0.54, 0.86) | < 0.001 | 0.75 (0.60, 0.93) | 0.01 |
| R635 | 0.78 (0.65, 0.93) | 0.008 | 0.80 (0.67, 0.95) | 0.01 |
| R425 | 0.84 (0.67, 1.06) | 0.15 | 0.94 (0.76, 1.17) | 0.62 |
| R270 | 0.86 (0.72, 1.03) | 0.11 | 0.91 (0.77, 1.08) | 0.28 |
| NR0 | 0.95 (0.77, 1.17) | 0.64 | 1.04 (0.86, 1.28) | 0.63 |
