## Supplementary Table 6 for "Dietary sugar exposure in early life and risk of adult mental health disorders: UK Biobank cohort study"

Supplementary table 6. Robustness check (Subset of population with later-life sugar intake, n = 21,221)

|  | Anxiety | | Depression | |
| --- | --- | --- | --- | --- |
| Group | HR (95% CI) | Adjusted P-value | HR (95% CI) | Adjusted P-value |
| Model 4 (additionally adjusted with sugar intake) | | | | |
| R1000 | 0.63 (0.44, 0.89) | 0.01 | 0.83 (0.58, 1.18) | 0.31 |
| R817 | 0.77 (0.48, 1.21) | 0.25 | 0.85 (0.55, 1.32) | 0.48 |
| R635 | 0.75 (0.53, 1.07) | 0.11 | 0.83 (0.58, 1.18) | 0.31 |
| R425 | 0.98 (0.58, 1.42) | 0.67 | 1.06 (0.69, 1.65) | 0.77 |
| R270 | 0.82 (0.57, 0.90) | 0.28 | 1.08 (0.76, 1.54) | 0.64 |
| NR0 | 1.00 (0.66, 1.52) | 0.98 | 1.23 (0.82, 1.86) | 0.30 |
