## Supplementary Table 7 for "Dietary sugar exposure in early life and risk of adult mental health disorders: UK Biobank cohort study"

Supplementary table 7. Significant ANCOVA brain region

| Region | Levene P_Value | P_Value | P_Adjusted |
| --- | --- | --- | --- |
| Volume of grey matter in Amygdala (left) | 0.196172 | 1.06E-06 | 5.92E-06 |
| Volume of grey matter in Amygdala (right) | 0.40611 | 6.18E-09 | 1.02E-07 |
| Volume of grey matter in Angular Gyrus (right) | 0.384626 | 0.001463 | 0.003467 |
| Volume of grey matter in Brain-Stem | 0.977272 | 3.04E-19 | 3.89E-17 |
| Volume of grey matter in Central Opercular Cortex (left) | 0.109733 | 2.29E-08 | 2.63E-07 |
| Volume of grey matter in Central Opercular Cortex (right) | 0.303569 | 8.00E-08 | 6.83E-07 |
| Volume of grey matter in Crus I Cerebellum (left) | 0.559784 | 1.31E-07 | 9.30E-07 |
| Volume of grey matter in Crus I Cerebellum (right) | 0.176573 | 1.10E-07 | 8.30E-07 |
| Volume of grey matter in Crus II Cerebellum (left) | 0.324914 | 2.64E-05 | 9.94E-05 |
| Volume of grey matter in Crus II Cerebellum (right) | 0.398157 | 9.36E-06 | 4.05E-05 |
| Volume of grey matter in Cuneal Cortex (left) | 0.570099 | 0.018833 | 0.031307 |
| Volume of grey matter in Cuneal Cortex (right) | 0.610596 | 8.54E-05 | 0.000273 |
| Volume of grey matter in Frontal Medial Cortex (left) | 0.679627 | 0.002075 | 0.004742 |
| Volume of grey matter in Frontal Medial Cortex (right) | 0.889294 | 0.006671 | 0.013915 |
| Volume of grey matter in Frontal Operculum Cortex (left) | 0.917142 | 0.006849 | 0.013915 |
| Volume of grey matter in Frontal Operculum Cortex (right) | 0.316136 | 0.019404 | 0.031842 |
| Volume of grey matter in Frontal Orbital Cortex (left) | 0.979933 | 8.64E-06 | 3.95E-05 |
| Volume of grey matter in Frontal Orbital Cortex (right) | 0.675484 | 0.001434 | 0.003463 |
| Volume of grey matter in Frontal Pole (left) | 0.10173 | 2.13E-12 | 9.09E-11 |
| Volume of grey matter in Frontal Pole (right) | 0.543189 | 2.45E-07 | 1.57E-06 |
| Volume of grey matter in Heschl's Gyrus (includes H1 and H2) (left) | 0.479117 | 3.42E-08 | 3.36E-07 |
| Volume of grey matter in Heschl's Gyrus (includes H1 and H2) (right) | 0.440078 | 9.25E-08 | 7.40E-07 |
| Volume of grey matter in I-IV Cerebellum (left) | 0.883379 | 0.008189 | 0.016378 |
| Volume of grey matter in I-IV Cerebellum (right) | 0.44548 | 0.000266 | 0.000774 |
| Volume of grey matter in IX Cerebellum (left) | 0.703864 | 0.012672 | 0.022846 |
| Volume of grey matter in IX Cerebellum (right) | 0.889464 | 0.005738 | 0.012449 |
| Volume of grey matter in IX Cerebellum (vermis) | 0.70438 | 0.011512 | 0.021992 |
| Volume of grey matter in Inferior Frontal Gyrus, pars opercularis (left) | 0.753245 | 0.009228 | 0.017897 |
| Volume of grey matter in Inferior Temporal Gyrus, posterior division (left) | 0.225895 | 0.01194 | 0.022149 |
| Volume of grey matter in Inferior Temporal Gyrus, posterior division (right) | 0.58492 | 0.023017 | 0.037294 |
| Volume of grey matter in Insular Cortex (right) | 0.102677 | 0.027792 | 0.044468 |
| Volume of grey matter in Juxtapositional Lobule Cortex (formerly Supplementary Motor Cortex) (left) | 0.176051 | 0.014018 | 0.024351 |
| Volume of grey matter in Lateral Occipital Cortex, superior division (left) | 0.980446 | 3.50E-05 | 0.000128 |
| Volume of grey matter in Lateral Occipital Cortex, superior division (right) | 0.857872 | 8.69E-09 | 1.24E-07 |
| Volume of grey matter in Lingual Gyrus (left) | 0.778758 | 0.018465 | 0.031099 |
| Volume of grey matter in Lingual Gyrus (right) | 0.427956 | 0.000602 | 0.001676 |
| Volume of grey matter in Middle Temporal Gyrus, posterior division (right) | 0.427566 | 0.00628 | 0.013397 |
| Volume of grey matter in Occipital Fusiform Gyrus (left) | 0.098481 | 0.000155 | 0.000472 |
| Volume of grey matter in Occipital Fusiform Gyrus (right) | 0.18727 | 3.81E-05 | 0.000135 |
| Volume of grey matter in Occipital Pole (left) | 0.538686 | 1.13E-06 | 6.04E-06 |
| Volume of grey matter in Occipital Pole (right) | 0.059361 | 8.38E-05 | 0.000273 |
| Volume of grey matter in Paracingulate Gyrus (right) | 0.771735 | 3.90E-11 | 1.25E-09 |
| Volume of grey matter in Parahippocampal Gyrus, posterior division (left) | 0.284184 | 9.81E-06 | 4.05E-05 |
| Volume of grey matter in Parahippocampal Gyrus, posterior division (right) | 0.214802 | 0.006799 | 0.013915 |
| Volume of grey matter in Planum Polare (left) | 0.488653 | 2.33E-07 | 1.57E-06 |
| Volume of grey matter in Planum Temporale (left) | 0.090796 | 0.000766 | 0.002002 |
| Volume of grey matter in Postcentral Gyrus (left) | 0.79296 | 0.000232 | 0.00069 |
| Volume of grey matter in Postcentral Gyrus (right) | 0.132481 | 0.00381 | 0.008409 |
| Volume of grey matter in Precentral Gyrus (left) | 0.696444 | 0.002073 | 0.004742 |
| Volume of grey matter in Precentral Gyrus (right) | 0.76616 | 2.33E-05 | 9.03E-05 |
| Volume of grey matter in Precuneous Cortex (left) | 0.667627 | 0.00066 | 0.001796 |
| Volume of grey matter in Precuneous Cortex (right) | 0.882028 | 6.83E-05 | 0.00023 |
| Volume of grey matter in Subcallosal Cortex (left) | 0.460977 | 0.014078 | 0.024351 |
| Volume of grey matter in Superior Frontal Gyrus (left) | 0.550013 | 0.014983 | 0.02557 |
| Volume of grey matter in Superior Temporal Gyrus, anterior division (left) | 0.330678 | 0.000986 | 0.002474 |
| Volume of grey matter in Superior Temporal Gyrus, posterior division (left) | 0.72061 | 0.012234 | 0.02237 |
| Volume of grey matter in Superior Temporal Gyrus, posterior division (right) | 0.678659 | 0.003733 | 0.008383 |
| Volume of grey matter in Temporal Fusiform Cortex, anterior division (left) | 0.122517 | 7.98E-06 | 3.78E-05 |
| Volume of grey matter in Temporal Fusiform Cortex, posterior division (left) | 0.800806 | 0.000141 | 0.00044 |
| Volume of grey matter in Temporal Fusiform Cortex, posterior division (right) | 0.364551 | 0.011937 | 0.022149 |
| Volume of grey matter in Temporal Occipital Fusiform Cortex (right) | 0.967652 | 0.008598 | 0.016932 |
| Volume of grey matter in V Cerebellum (left) | 0.556173 | 0.000767 | 0.002002 |
| Volume of grey matter in V Cerebellum (right) | 0.722759 | 0.001316 | 0.00324 |
| Volume of grey matter in VI Cerebellum (left) | 0.277854 | 2.27E-09 | 5.65E-08 |
| Volume of grey matter in VI Cerebellum (right) | 0.786864 | 8.97E-13 | 5.74E-11 |
| Volume of grey matter in VI Cerebellum (vermis) | 0.901526 | 0.000448 | 0.001274 |
| Volume of grey matter in VIIIa Cerebellum (left) | 0.635236 | 3.42E-07 | 2.08E-06 |
| Volume of grey matter in VIIIa Cerebellum (right) | 0.658068 | 2.65E-09 | 5.65E-08 |
| Volume of grey matter in VIIIa Cerebellum (vermis) | 0.963784 | 1.41E-06 | 7.22E-06 |
| Volume of grey matter in VIIIb Cerebellum (left) | 0.568863 | 0.013645 | 0.024259 |
| Volume of grey matter in VIIIb Cerebellum (right) | 0.693232 | 0.000872 | 0.002232 |
| Volume of grey matter in VIIIb Cerebellum (vermis) | 0.415376 | 5.54E-05 | 0.000192 |
| Volume of grey matter in VIIb Cerebellum (left) | 0.908556 | 6.38E-09 | 1.02E-07 |
| Volume of grey matter in VIIb Cerebellum (right) | 0.745842 | 1.17E-08 | 1.50E-07 |
| Volume of grey matter in VIIb Cerebellum (vermis) | 0.748021 | 1.80E-05 | 7.19E-05 |
| Volume of grey matter in Ventral Striatum (left) | 0.052903 | 6.69E-08 | 6.12E-07 |
| Volume of grey matter in Ventral Striatum (right) | 0.221939 | 9.63E-06 | 4.05E-05 |
| Volume of grey matter in X Cerebellum (left) | 0.810092 | 8.66E-07 | 5.04E-06 |
| Volume of grey matter in X Cerebellum (vermis) | 0.337363 | 2.46E-08 | 2.63E-07 |
| Volume of grey matter in X Cerebellum (right) | 0.45451 | 5.45E-06 | 2.68E-05 |
