## Supplementary Table 8 for "Dietary sugar exposure in early life and risk of adult mental health disorders: UK Biobank cohort study"

Supplementary Table 8. The loadings of the latent variables in SEM models (Wald tests, two-sided P values)

| Latent Variables | Estimate | Z value | P(>\|z\|) |
| --- | --- | --- | --- |
| **Sweet taset liking =~** |  |  |  |
| Liking for biscuits | 1.00 |  |  |
| Liking for cake | 1.11 | 37.201 |  |
| Liking for cake icing | 0.87 | 24.503 | 0.000 |
| Liking for croissant | 0.697 | 22.524 | 0.000 |
| Liking for dark chocolate | 0.182 | 7.330 | 0.000 |
| Liking for cheesecake | 0.819 | 26.672 | 0.000 |
| Liking for ice cream | 0.789 | 28.268 | 0.000 |
| Liking for jam | 0.834 | 29.202 | 0.000 |
| Liking for marzipan | 0.497 | 16.076 | 0.000 |
| Liking for milk chocolate | 0.746 | 26.123 | 0.000 |
| Liking for sweet foods | 1.032 | 33.258 | 0.000 |
| **Sweet intake =~** |  |  |  |
| Free sugar | 1.00 |  |  |
| Total sugar | 0.692 | 10.592 | 0.000 |
| **Brain MRI =~** |  |  |  |
| Brain stem | 1.00 |  |  |
| Crus I Cerebellum (left) | 1.255 | 28.079 | 0.000 |
| Crus I Cerebellum (right) | 1.344 | 30.155 | 0.000 |
| Crus II Cerebellum (right) | 1.655 | 34.715 | 0.000 |
| Occipital Fusiform Gyrus (right) | 0.875 | 20.951 | 0.000 |
| VI Cerebellum (left) | 1.762 | 35.024 | 0.000 |
| VI Cerebellum (right) | 1.759 | 35.171 | 0.000 |
| VIIIa Cerebellum (left) | 1.902 | 35.933 | 0.000 |
| VIIIa Cerebellum (right) | 1.799 | 33.796 | 0.000 |
| VIIb Cerebellum (right) | 1.887 | 36.196 | 0.000 |
| VIIb Cerebellum (left) | 1.930 | 36.096 | 0.000 |
